# Identification of HBV and HCV transmission in multi-transfused Thalassemia patients of Azad Jammu and Kashmir, Pakistan

**DOI:** 10.1101/2022.01.25.22269811

**Authors:** Majid Mahmood, Marum Aslam, Nasib Zaman, Fahed Parvaiz, Ali Muhammad, Nausheen Irshad, KAamir Pervez, Khawaja Shafique Ahmad

**Affiliations:** Department of Zoology, University of Poonch Rawalakot, AJ&K, Pakistan; Institute of Microbiology and Biotechnology, University of Swat, Swat, Pakistan; Department of Biosciences, COMSATS University, Islamabad, Pakistan; Department of Botany, University of Poonch Rawalakot, AJ&K, Pakistan

**Keywords:** Thalassemia, Hepatitis B virus, Hepatitis C virus, Blood transfusion, Azad Jammu and Kashmir

## Abstract

**Background:** Thalassemia patients need regular blood transfusion which saves their lives but increases the risk of acquiring viral infections like hepatitis B virus (HBV), hepatitis C virus (HCV) and human immunodeficiency virus (HIV).

**Objective(s):** This study aimed to screen out the HCV and HBV infections in Thalassemia patients receiving regular blood in three centers of Azad Jammu and Kashmir (AJK), Pakistan.

**Materials and Methods:** It was a cross sectional, multicenter study involving 224 Thalassemia patients. The study was carried out from August 2019 to January 2020 in Thalassemia centers of Sheikh Khalifa Bin Zaid hospital Rawalakot, District Headquarters hospital (DHQ) Palandari, and Combined Military Hospital (CMH) Muzaffarabad, AJK. A total of 224 patients were screened out for HCV and HBV. Indirect ELISA was carried out to detect anti HCV antibodies while direct ELISA was carried out to detect the HBsAg from all the samples using commercially available ELISA kits.

**Results:** Overall, 12 patients (5.36%) were found to be positive for HBV while 63 (28.1%) were positive for HCV. HBV positive patients included 7 (5.69%) males and 5 (4.95%) females while HCV positive patients included 26 (41.27%) males and 37 (58.73%) female patients. Both the viruses were more common among elder patients as compared to younger patients.

**Conclusion:** Prevalence of HCV is very high and that of HBV is moderately high among Thalassemia patients of AJK.

## Introduction

Hepatitis B virus (HBV) and hepatitis C virus (HCV) are currently two of the major killers of human being all over the world. It is estimated that about 400 million people are infected with HBV and 200 million with HCV all around the globe [1-3]. HBV and HCV are highly prevalent in Pakistan as about 10 million people are infected with HBV and another 10 million are infected with HCV [4-5]. Both of the HBV and HCV are blood borne viruses and transmitted through blood transfusion, injections, surgical instruments, dental procedures, piercing, tattooing, tooth brushes, razors, pedicuring equipments, menicuring equipments and other invasive practices. Moreover, it has also been reported that both the viruses can be transmitted from infected mother to newborn which is called vertical transmission while HBV is also transmitted through sexual contacts [6-9]. Both the HBV and HCV cause liver damage. They lead to chronic liver disease, cirrhosis, hepatocellular carcinoma (HCC), end stage liver disease and death and are the most common transfusion transmitted infections (5, 10-11).

Thalasseamia is a group of genetic disorders which is majorly divided into alpha-thalassemia and beta thalassemia [12]. Beta-thalassemia is the most prevalent genetic problem worldwide and its carrier rate in Pakistan is about 5-7% [13].This is an autosomal recessive disorder characterized by reduced or absent beta globin chain [12].

Thalassemia patients need regular blood transfusion for maintaining their hemoglobin level [14].Regular blood transfusion in these patients saves their lives but increases the risk of acquiring viral infections like hepatitis B virus (HBV), hepatitis C virus (HCV) and human immunodeficiency virus (HIV) (14-15). So, these patients have a great risk of obtaining HCV, HBV, HIV and other infections as compared to the normal population [14-15]. Although, the blood screening before transfusion is a normal practice now, still the transmission of viruses occurs even from healthy blood donors. Moreover, the patients sometimes receive blood from professional blood donors when the blood is needed urgently [16-17].

The aim of current study was to find out the prevalence of both HCV and HBV in blood receiving thalassemia patients from three thalassemia centers of AJK, as enough scientific literature is not available on topic from the region of AJK, Pakistan.

## Patients and Methods

### Study Duration, Design and Settings

This was a multicenter cross sectional study carried out from August 2019 to January 2020 in Thalassemia centers of Sheikh Khalifa Bin Zaid hospital Rawalakot, District Headquarters hospital (DHQ) Palandari, and Combined Military Hospital (CMH) Muzaffarabad, AJK.

### Inclusion and Exclusion Criteria

Thalassemia patients who were receiving blood in any of the selected hospitals having no known HCV or HBV infection in their family were included in the study.

### Patients Enrolment

A total of 224 thalassemia patients, regularly receiving blood in the selected hospitals, who met the inclusion criteria, were enrolled in the study. Out of these, 81 were enrolled in Sheikh Khalifa Bin Zaid hospital Rawalakot, 73 in District Headquarters hospital Palandari and 70 in Combined Military Hospital, Muzaffarabad. The patients included 123 (54.9%) male and 101 (45.1%) female with age ranging from 1 to 18 years.

### Patient’s Data Collection

The relevant data of risk factors was collected on a prescribed proforma. The factors included: patient’s blood group, age, gender, period of blood transfusion, source of blood, parent’s relationship, and cast of the patient.

### Sample Collection and Screening

Blood samples were taken with the help of 5 ml sterile syringes in EDTA coated tubes from all patients. The serum was separated by centrifugation at 3000 rpm for five minutes and stored at -20^°^C for further analysis. Blood samples of all the patients were subjected to Enzyme Linked Immuno Sorbant Essay (ELISA). Indirect ELISA was carried out to detect anti-HCV antibodies while direct ELISA was carried out to detect HBsAg using commercially available ELISA kits following the manufacturers protocol (Genway Biotech, United Kingdom).

### Statistical Analysis

All the data was entered and analyzed in Statistical Package for Social Sciences (SPSS) version 16.0. Chi square test was applied for qualitative comparisons where needed.

### Ethical Approval and Informed Consent

The study was approved by “human and animals ethics committee”, university of Poonch Rawalakot AJK. An informed consent form was filled and signed by each participant before the start of blood screening.

## Results

### General Characteristics of Patients

Out of the total 224 Thalassemia patients, 123 (54.9%) were male and 101 (45.1%) were female. The most common blood group was B which was detected in 101 patients (45.1%) while 64 (28.6%) patients had O blood group, 43 (19.2%) had A and 16 (7.14%) patients had AB blood group. Majority of the patients were Rh positive (90.6%) while a few were Rh negative (Table 1). Cousin marriage was traced in the parents of 78.57% of total cases while in remaining 21.43% of the cases, an unrelated parents’ marriage was traced.

**Table1.**
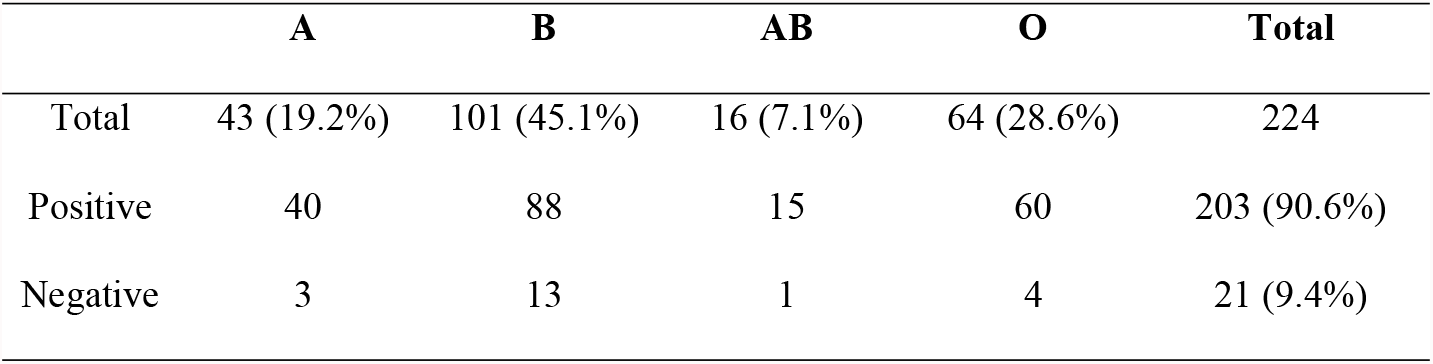
Distribution of blood group and Rh antigen among the study patients

|  | **A** | **B** | **AB** | **O** | **Total** |
| --- | --- | --- | --- | --- | --- |
| Total | 43 (19.2%) | 101 (45.1%) | 16 (7.1%) | 64 (28.6%) | 224 |
| Positive | 40 | 88 | 15 | 60 | 203 (90.6%) |
| Negative | 3 | 13 | 1 | 4 | 21 (9.4%) |

### Overall and Center Wise Prevalence

Out of 224 total patients, 81 were registered in Sheikh Khalifa Bin Zaid Hospital, Rawalakot, 73 in District Headquarters Hospital Palandari, and 70 in Combined Military Hospital (CMH), Muzaffarabad. Overall, 12 patients (5.36 %) were found to be positive for HBV while 63 (28.1%) were positive for HCV (Table 2).

**Table 2.** Comparison of nfection prevalence among three study centers and between male and female

|  |  | Total (n=224) | Rawalakot (n=81) | Palandari (n=73) | Muzaffarabad (n=70) | Significance |
| --- | --- | --- | --- | --- | --- | --- |
| HBV | Total (n=224) | 12 (5.36%) | 3 (3.70%) | 0 | 9 (12.86%) | 0.002 |
|  | Male (n=123) | 7 (5.7%) | 2 | 0 | 5 |  |
|  | Female (n=101) | 5 (4.95%) | 1 | 0 | 4 |  |
|  | Significance | 0.783 |  |  |  |  |
| HCV | Total (n=224) | 63 (28.12%) | 14 (17.28%) | 21 (28.77%) | 28 (40.0%) | 0.008 |
|  | Male (n=123) | 26 (21.13%) | 5 | 10 | 11 |  |
|  | Female (n=101) | 37 (36.63%) | 9 | 11 | 17 |  |
|  | Significance | 0.000 |  |  |  |  |

Among 81 patients in Sheikh Khalifa Bin Zaid Al-Nahyan Hospital Rawalakot, 14 (17.28%) were positive for HCV including 5 male and 9 female patients, while only 3 (3.7%) patients were HBV positive including 2 males and 1 female. In District Headquarters Hospital Palandari, there was no HBV positive patient while 21 patients (28.76%) were HCV positive including 10 (47.62%) male and 11 (52.38%) female. In Combined Military Hospital Muzaffarabad, 9 patients (12.86%) were HBV positive including 5 males and 4 females while 28 (40%) were positive for HCV including 11 male and 17 female patients. The prevalence of HBV was highest in the patients of Muzaffarabad as compared to the patients in Rawalakot and the difference was statistically significant (*P = 0*.*002*) while there was no positive case of HBV in Palandari center (Table 2).

The highest number of HCV positive patients was found in Muzaffarabad where 28 (40%) out of 70 were positive. It was followed by Palandari, with 21 (28.76%) out of 73 positive patients while the lowest percentage was found in Rawalakot with 14 (17.28%) out of 81 positive patients. There was a significant difference of percentage prevalence among three areas for HCV (*P=0*.*008*).

Out of the positive patients, 7 males (5.69%) and 5 females (4.95%) were positive for HBV while 26 (21.14%) males and 37 (36.63 %) females were positive for HCV. There was no significant difference of HBV infection between male and female, while the prevalence of HCV was significantly higher (*P=0*.*008*) in female patients as compared to male patients (Table 2).

### Age wise Prevalence

In present study, the mean age of patients was calculated to be 7.51± 4.67 years, ranging from 1-18 years. The patients were divided into three age groups. The first group comprised of the patients of 1 to 6 years and there were 118 patients in this age group. Out of these 118 patients, 3 (2.54%) were positive for HBV and all were present in Muzaffarabad center while 16 (13.56%) were positive for HCV which included 2 (1.7%) in Rawalakot, 5 (4.2%) in Palandari and 9 (7.6%) in Muzaffarabad center (Table 3).

**Table 3.** Comparison of infection prevalence among different age groups of the patients

|  |  | **1-6 years (n=118)** | **7-12 years (n=72)** | **13-18 years (n=34)** | ***Significance*** |
| --- | --- | --- | --- | --- | --- |
| **HBV** | *Total* | 3 (2.54%) | 6 (8.33%) | 3 (8.82% ) | ***0.038*** |
|  | *Rawalakot (n=81)* | 0 | 2 | 1 |  |
|  | *Palandari (n=73)* | 0 | 0 | 0 |  |
|  | *Muzaffarabad (n=70)* | 3 | 4 | 2 |  |
| **HCV** | *Total* | 16 (13.56%) | 23 (31.9%) | 23 (67.6%) | ***0.000*** |
|  | *Rawalakot (n=81)* | 2 (1.7%) | 7 (9.7%) | 5 (14.7%) |  |
|  | *Palandari (n=73)* | 5 (4.2%) | 8 (11.1%) | 8 (23.5%) |  |
|  | *Muzaffarabad (n=70)* | 9 (7.6%) | 8 (11.1%) | 11 (32.35%) |  |

Second group included patients from 7 to 12 years in which 72 patients were present. Out these 72 patients, 6 (8.33 %) were positive for HBV including 2 (2.78%) in Rawalakot and 4 (5.55%) in Muzaffarabad while 23 (31.94%) were positive for HCV including 7 (9.7%) in Rawalakot center, 8 (11.1%) in Palandari and 8 (11.1%) in Muzaffarabad center (Table 3).

Third age group comprised of 13 to 18 years which included 34 patients. In this group, 3 (8.82%) were positive for HBV including 1 (2.94%) in Rawalakot and 2 (5.88%) in Muzaffarabad while 23 (67.64%) were positive for HCV including 5 (14.7%) in Rawalakot, 8 (23.5%) in Palandari and 11(32.35%) in Muzaffarabad center. There was a significant difference of both HCV and HBV infection (*P < 0*.*05*) among the age groups as the older age patients had significantly higher percentage of both infections as compared to younger patients.

## Discussion

Blood transfusion is the important risk factor for the transmission of viruses like HIV, HBV and HCV. Thalassemia patients require regular blood transfusion for survival and have always a high risk of getting these infections. The present study was aimed to calculate the prevalence of HBV and HCV infections in three thalassemia centers of the state of Azad Jammu and Kashmir. The study included 224 thalassemia patients who were screened for hepatitis C and B viruses. The prevalence of these viruses was found to be 28.1% and 5.36% respectively. Overall prevalence of HCV was higher than that of HBV. Almost similar results were also reported previously in a study from Pakistan who reported HCV in 29% and HBV in 8% of the thalassemia patients [18]. Another study conducted in Pakistan reported that 39% of thalassemia patients are infected with HCV and 5% with HBV [19]. However, a very high prevalence of HCV among thalassemia patients in Pakistan (55%) was reported in another earlier study [20]. So, the thalassemia patients in Pakistan are highly infected with HCV and moderately with HBV.

On the base of present study and previous literature, it is observed that infection rate of HCV and HBV in thalassemia patients is very high in Pakistan and AJK when compared with some of the nearby countries like Iran, Iraq and Palestine, as the infection percentage of HCV and HBV in thalassemia patients was reported to be 3.8% and 3.8% respectively in Iran, 3.8% and 2.5% respectively in Iraq, and 10% and <1% respectively in Palestine [21-23]. The fact that thalassemia patients of Pakistan and AJK have very high HCV infection and a moderate HBV infections may be linked with the fact that prevalence of HCV is already high and that of HBV is also moderate to high in general population of Pakistan and AJK [24-26]. The prevalence of both HCV and HBV is very high in Pakistan when compared to advanced countries like Germany where the prevalence of both viruses ranges from 0.2% to 1.9% [27]. So, the prevalence of HCV and HBV in thalassemia patients of the region corresponds to the prevalence of these infections in general population. However, it is much higher than the prevalence in general population which seems because of the high risk posed by multiple blood transfusions to these patients and lack of standard blood screening system prior to transfusion. This fact is also supported by the observation that the patients with more transfusions have more prevalence than those with less transfusions received (Table 3).

Overpopulation, poverty and lack of resources are some more factors possibly contributing towards the high prevalence of HCV and HBV in thalassemia patients of the country which limit the blood screening tests to low price only, and in turn, allow the viral infections to pass along with blood to the patients. Besides these, another fact that matters reasonably is the selection of blood donors. In case of thalassemia patients who often need blood urgently, the donor is not always been selected by some special criteria, but the blood is received from anyone ready to donate or even sometime is purchased from professional blood donors who may be injectable drug users (IDUs). Collectively, because of low resources and shorter time, the proper and up-to the standards screening may not be possible in the region to control efficiently the transfusion transmitted viruses.

The prevalence of HCV and HBV was found to be significantly higher in older age patients as compared to those of younger age (Table 3). This finding may be associated with the fact that the older patients have lived more age with thalassemia and received more blood transfusions than the younger ones. As, higher the number of transfusion more will be the exposure to viruses and risk of infection, the elder patients are likely to have more infection than the younger patients who received less blood. Similar findings and reasons have been previously reported too [22, 26].

The highest prevalence of HCV was found in Muzaffarabad followed by Palandari and then Rawalakot. The prevalence of HBV was also higher in the patients of Muzaffarabad center as compared to the patients in Rawalakot and Palandari centers. The center wise differences of HCV and HBV prevalence may be because of the local differences of HBV and HCV prevalence in these areas or may be because of some differences in screening procedure or because of some unknown reasons. Currently, no study or other published literature is available to confirm any fact or reason of these differences. More studies may provide some better explanation.

Current research shows the alarming situation of HCV and HBV transmission in thalassemia patients through blood transfusion in the area of AJK, Pakistan. The results will guide the healthcare professionals and the thalassemia patients to screen the blood before transfusion using more efficient methods. It may influence the policy of blood transfusion in the area and lead to hepatitis control strategies.

## Conclusions

The current study concludes that HCV infection is very high (28.1%) while HBV is moderately high (5.36%) in thalassemia patients of the region of Azad Jammu and Kashmir, Pakistan. The prevalence is higher as compared to the prevalence of these viruses in other countries. Moreover, the prevalence of the viruses is higher in older age patients as compared to younger patients.

## Data Availability

All relevant data are within the manuscript and its Supporting Information files.

No

## Acknowledgment

Miss Saba Nosheen and Miss Sumayya Siddique are acknowledged for their role in data collection.

